# Parsing inter-individual variability in the digital phenotype across the menstrual cycle

**DOI:** 10.64898/2026.08.26.26361403

**Authors:** Loran Knol, Anisha Nagpal, Faraz Hussain, Christian F. Beckmann, Alex Leow, Tory A. Eisenlohr-Moul, Andre F. Marquand

## Abstract

Digital phenotyping, which is defined as quantifying someone’s behaviour with digital devices, provides unprecedented opportunities for understanding human mental health but is hampered by high levels of inter-individual variability. Here, we propose a new method to address this, parsing inter-individual variability by decomposing the digital phenotype dynamics into latent trajectories and using each individual’s trajectory membership as a moderator when modelling psychopathology over the same timeframe. We applied our method in the context of mood symptom exacerbation across the menstrual cycle, where symptom severity and timing are inconsistent between individuals. Using the BiAffect platform to collect smartphone typing dynamics, we found stable trajectories in smartphone movement rate: one group of participants showed substantial movement rate fluctuations across the menstrual cycle, whilst the others did not. Participants with movement fluctuations displayed increased fluctuations across the cycle in prospective anhedonia and depression ratings, but not in anxiety, irritability, and suicidal ideation.

## Main

The continued digitisation of our environment provides unprecedented opportunities for understanding human mental health and behaviour^1,2^. Correspondingly, digital devices, ranging from wearables to smartphones, are being employed at scale to measure markers of mental health and cognition in the real world over years at a time, for example to predict relapse in schizophrenia^3^, progression in mild cognitive impairment and dementia^4^, and depressive and manic mood episodes^5^, among many other things^6^. Collectively, this is called digital phenotyping^7,8^. A key factor underlying its scalability across sample size and study duration is its potential for passive, unobtrusive, and ecologically valid measurements that require minimal user interaction.

Despite these promises, however, the translation of digital phenotyping toward clinical implementation has been slow^2^. One of the most important reasons is that the analysis of digital phenotypes is plagued by high inter-individual variability on several levels^9^, including variation in both behavioural manifestation of clinical conditions and device use patterns that are due to healthy variability and no consequence of any pathology. Together, this variability makes group comparisons challenging, but drawing individualised conclusions can be equally difficult without an adequate reference to compare an individual’s data against. Moreover, digital phenotyping data often display non-linear and possibly rhythmic dynamics over time. This occurs, for example, in disorders featuring distinct or recurrent episodes (e.g., bipolar disorder) or in sleep^10,11^. There is therefore a need for modelling approaches that can dissect different cluster profiles of non-linear variation related to symptoms on the basis of temporal dynamics in the data. Parsing this variability and relating it to psychopathology would increase the potential of digital phenotyping to function as a passive marker of psychopathology, ultimately facilitating its use for precision medicine approaches in which such markers could be used to draw individualised conclusions and inform diagnosis and treatment^1^.

In this study, we propose a method to address these issues. We identify latent trajectories in digital phenotypes and relate these to meaningful differences in psychological symptom trajectories. Our approach consists of a three-stage procedure: first, we characterise baseline dynamics in the digital phenotype. Next, we parse heterogeneity in the digital phenotype dynamics by decomposing them into latent trajectories^12^. Then, finally, we model psychopathology trajectories across time, in which the digital phenotype trajectory membership moderates the relationship between time and symptom severity. An overview of our approach is given in Figure 1A.

**Figure 1:**
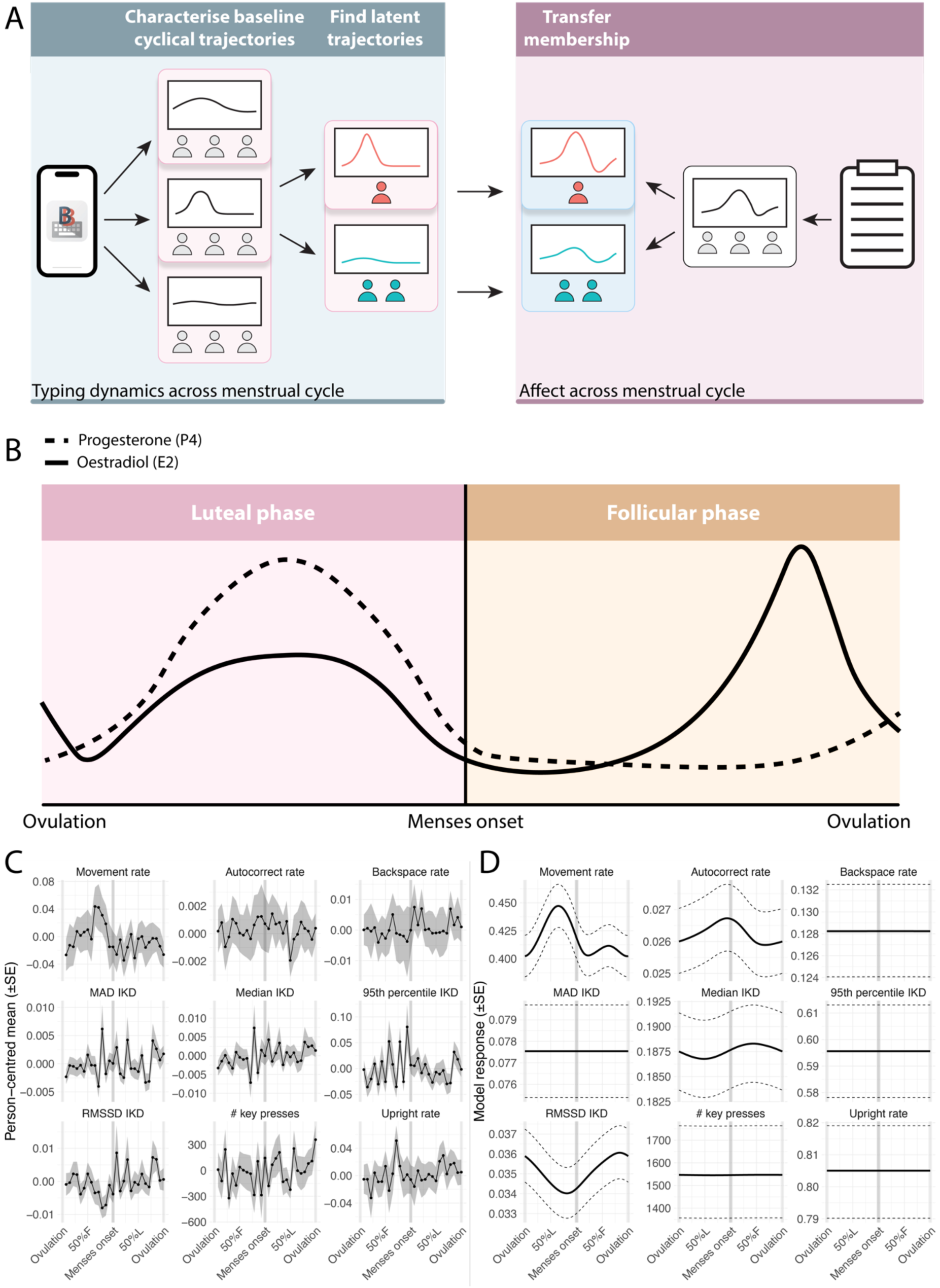
Overview of study approach and baseline results of typing dynamics over the menstrual cycle. **A** Illustration of analysis steps. We first characterise menstrual cycle trajectories in the smartphone keyboard dynamics collected by BiAffect. Second, we use mixture models to discover subtrajectories in those BiAffect metrics that exhibit significant non-zero baseline trajectories, recording which participants were assigned to which latent trajectory. Third, we transfer these trajectory memberships to the psychopathology domain, where different curves are fitted for each of the latent trajectory groups discovered in the keyboard data. **B** Illustration of sex hormone flux across the menstrual cycle. Progesterone and oestradiol peak in the luteal phase and then start declining, triggering menses. Oestradiol peaks again in the late-follicular phase, just before ovulation. **C** Participant-centred means of their keyboard dynamics across the cycle. Weighted means and Kish’s design effect (for standard errors) were used for all rate metrics as they consisted of weighted observations. **D** Response means for all BiAffect metrics as estimated by GAMMs. MAD = Median absolute deviation; IKD = Inter-key delay; RMSSD = Root mean square of successive differences.

We explore this issue in the formidably challenging context of mood symptom exacerbation across the menstrual cycle^13^. We utilise data from the transdiagnostic, pre-registered randomised crossover trial CLEAR-3 (NCT04112368), which investigated the hormonal mechanisms behind perimenstrual exacerbation of suicidal ideation in a sample recruited for natural cycling and suicidality. In a substantial number of cases, ovulating individuals experience cyclical increases in psychological symptom severity due to altered sensitivity to normal ovarian steroid fluctuations^14,15^, but with considerable between-person differences in symptom timing^16^. The accurate identification of cyclical steroid sensitivity requires prospective daily ratings over an extended timeframe^17^, which is highly burdensome and unwieldy^18^. Therefore, to aid the diagnosis of these disorders with passive measures of smartphone interactions, we use BiAffect, a well-validated iOS app that measures typing dynamics by replacing the default iPhone keyboard with one that collects data on metrics such as typing speed, phone orientation, and phone movement. In other words, it measures how someone types, not what they type. This has been shown to be sensitive to many aspects of cognition and psychopathology^19–30^.

Our study was motivated by the clinical intuition that a subgroup of individuals would have amplified fluctuations in passive measures across the menstrual cycle that may help to identify individuals with cyclical patterns in the severity of their affective symptoms.

## Results

### Demographics

For the intervention and placebo arms of the CLEAR-3 trial, participants were recruited from a pool of individuals under outpatient mental healthcare and with past-month suicidal ideation (SI). They completed at least one month of baseline data collection, as well as a month of wash-out during cross-over. In addition, healthy controls were recruited for only the baseline period. We used baseline and, if available, washout data from both participant groups for the present study. After filtering (see Methods), baseline typing dynamics were analysed for 72 participants, out of which 59 had past-month SI and 13 were healthy controls. Together, they contributed 3574 observations of daily mood ratings and daily BiAffect aggregates (average: 49.6 observations per participant, range: 6-114). Due to slightly different data availability per typing metric, the actual number of participants and observations that the models were fitted with sometimes was slightly lower (Supplementary Table 2). Sample demographics are given in Supplementary Table 1.

### Baseline typing dynamics

As described above, we used BiAffect to passively collect digital biomarkers of cognition and mood. BiAffect collects data about the type and timing of individual key presses as well as the movement and orientation of the phone while typing. Most of these data are then preprocessed within typing sessions before being aggregated to the daily level. Typing sessions are initiated with the user’s first key press and terminated by either closing the keyboard or six seconds of inactivity. Preprocessing details can be found in the Methods, but to summarise, the BiAffect metrics used in this study were as follows: 1) median inter-key delay (IKD), an inverse representation of typing speed, 2) 95^th^ percentile IKD, a measure of pausing within typing sessions, 3) median absolute deviation (MAD) IKD, a measure of typing speed variability, 4) root mean square of successive differences (RMSSD) of the median IKD, another measure of typing speed variability that takes temporal order into account, 5) total daily key press count, 6) autocorrect rate, the daily number of autocorrects divided by the daily key press count 7) backspace rate, similarly defined as autocorrect rate, 8) percentage of typing sessions spent upright, and 9) percentage of typing sessions during which movement was recorded.

We used generalised additive mixed models (GAMMs) to model non-linear trajectories of typing data across the menstrual cycle. GAMMs are, in essence, generalised linear models with a sum of smooth functions of some covariates^31,32^. The smoothness of those curves, which prevents overfitting, is determined by a smoothing parameter and can be quantified by a curve’s effective degrees of freedom (edf). Larger edfs indicate less smooth (wigglier) curves, with an edf of 1 referring to a straight, sloped line.

For an overview of the phases and hormonal fluctuations of the menstrual cycle, see Figure 1B. Since the lengths of menstrual cycles and the timing of ovulation differ within and between individuals, with the follicular phase in particular showing large variability^33^, we used phase-aligned cycle time scaling (PACTS) to rescale all observed cycles to a common reference^34^. This approach uses menstruation and ovulation as anchor points for its scaling and avoids some of the drawbacks of counting methods, where the cycle phase is estimated by counting days forwards or backwards from menses. PACTS can be done in a menses- and in an ovulation-centred fashion. We report menses-centred results in the main text; for ovulation-centred results, see Supplementary Figure 4. For the baseline typing models, to model non-linearity across a cyclic domain we employed cyclic cubic regression splines as basis expansions of the menstrual cycle predictor. We additionally included participant-level random intercepts and slopes to account for the repeated-measures structure of the data^35^. P-values of the cycle effect were adjusted for a false discovery rate (FDR) of 0.05 across all nine models.

Response distributions and link functions were chosen to best match the response data type. We used scaled t-distributions with an identity link for all IKD metrics due to their heavy right tail (median IKD, MAD IKD, 95^th^ percentile IKD, and RMSSD IKD); a Tweedie distribution with a log link for count metrics (total number of key presses); and a quasibinomial distribution with a logit link for rate metrics, i.e. numbers in the interval [0, 1] with associated weights (movement rate, autocorrect rate, backspace rate, and upright rate).

Figure 1C shows participant-centred averages of the nine BiAffect metrics along with the mean responses estimated by the GAMMs. Between- and within-participant correlations across all metrics are given in Supplementary Figure 1. Whereas the participant-centred averages initially suggest some degree of cycle-related fluctuation for most metrics, this is not always reflected by the GAMMs, which is most likely due to the smoothing they enforce (Figure 1D). Broadly, movement rate shows a late-luteal increase and returns to its baseline shortly after menses (edf = 4.13, F = 12.49, p = 0.00011, p_adj_ = 0.0010). There seems to be a small secondary increase in the mid- to late-follicular phase. Typing metrics that showed a nominally significant cycle effect that did not survive FDR adjustment included autocorrect rate (edf = 2.25, F = 3.82, p = 0.041, p_adj_ = 0.092), log RMSSD IKD (edf = 1.71, F = 5.44, p = 0.035, p_adj_ = 0.092), and median IKD (edf = 1.66, F = 20.49, p = 0.012, p_adj_ = 0.053); see Supplementary Table 3 for all model estimates. Note that model residuals did not follow a normal distribution for log RMSSD IKD.

In what follows, we only show results for movement rate, as we expected robust effects of movement rate subgroups on daily symptom ratings based on previous work in the same sample^28^. In addition, movement rate showed the clearest subgroup structure out of all smartphone typing metrics. See Supplementary Figure 2 for results for the other typing metrics.

### Latent typing trajectories

To find subgroups in typing fluctuations across the menstrual cycle (i.e., latent trajectories), we adopted a growth mixture modelling (GMM) approach^36^, implemented here as a mixture of GAMMs. We refer to these GAMMs as the component models of the overarching mixture model. The mixture models had slightly more stringent filtering requirements than the baseline typing models. For movement rate while typing, they were built with data from 62 participants (52 with past-month SI and 10 healthy controls) contributing 3356 observations (average: 54.13 observations per participant, range: 11-114). Sample sizes for the other metrics for which we fitted mixture models are given in Supplementary Table 2.

Mixture models are often fitted using a two-step procedure that repeatedly alternates assigning observations to the component models with maximising the model likelihoods based on the assigned observations^37^. After convergence, this algorithm yields a list of probabilities indicating which component is most likely to have generated (a group of) observations; these probabilities are also known as responsibilities. Since our data is grouped by participants, we refer to these as participant responsibilities.

A challenge in this context is that GAMMs allocate a large part of model selection, i.e., selecting a suitable model flexibility, to the process that finds the smoothing parameter. These procedures have been designed for a single GAMM, and it is, at present, unclear how to adapt such procedures to mixtures of GAMMs. Ignoring this issue can lead to suboptimal mixture fits (Figure 2B, left panel). We therefore make several simplifying assumptions and do some of the model selection through out-of-sample validation.

**Figure 2:**
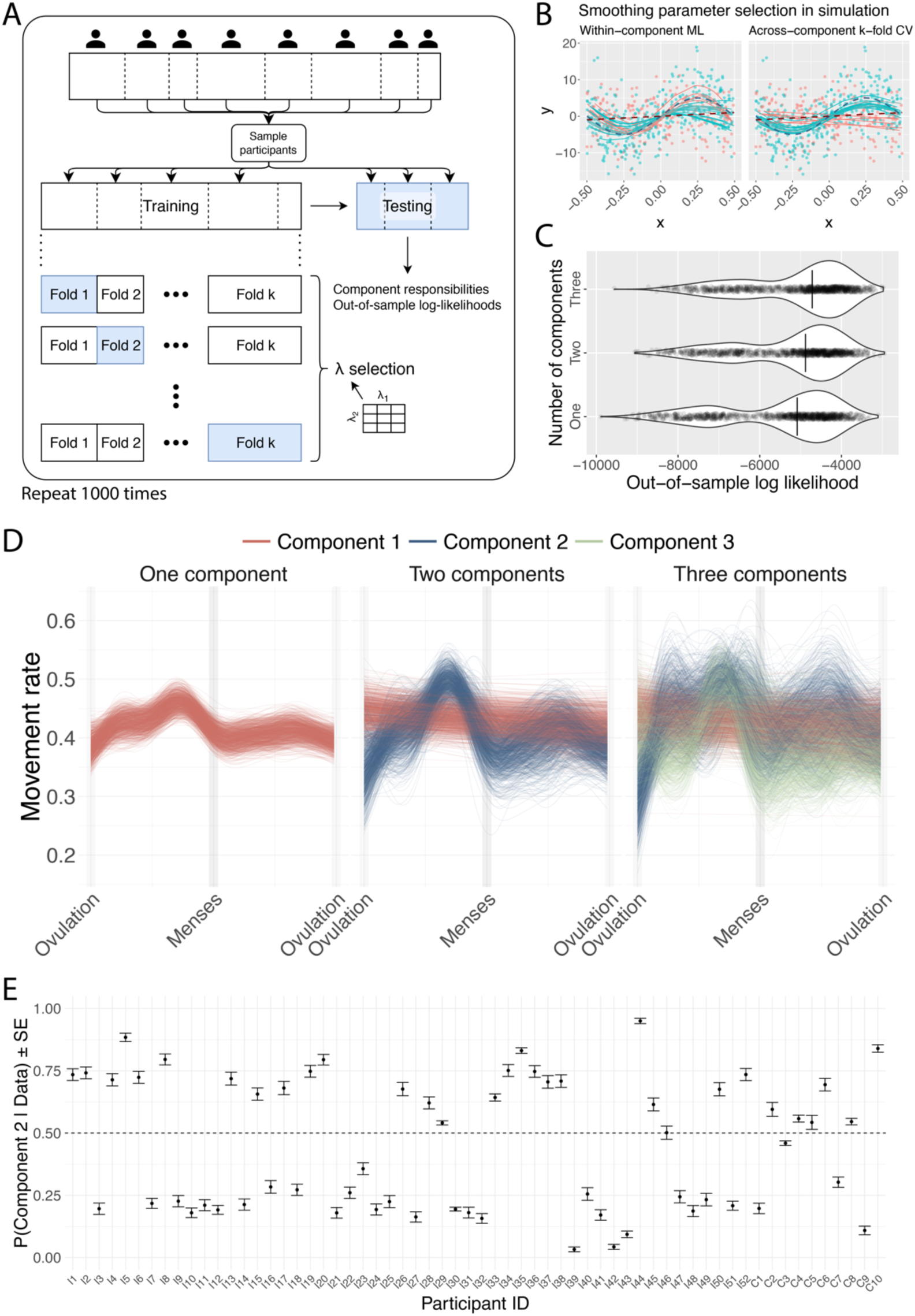
Mixture of GAMMs results. **A** Overview of the randomised CV procedure used to estimate out-of-sample log-likelihoods and participant-component responsibilities. Per iteration, participants are sampled into either the training or testing set. The mixture of GAMMs is trained, if need be, using k-fold CV to select the right smoothness parameters (λ). **B** Illustration of the benefit of doing smoothness selection across mixture components (right panel) rather than within (left panel) through a simulation. Dashed lines are group means, points are simulated data, and solid lines are fitted participant curves. Method in left panel fails to recover group with flat lines. **C** Violin plots of out-of-sample log-likelihoods for mixtures with one, two, and three components. Higher (i.e., less negative) is better. The vertical line in each violin plot indicates the sample median. **D** Line plots of individual GAMM fits, for all CV iterations. Because ordering of Components 2 and 3 was arbitrary for the three-component model, we did a principal component analysis of their spline coefficients for plotting purposes. We labelled those mixture components whose first principal component scores were above 0 as Component 2, and the rest as Component 3. **E** Mean Component 2 responsibilities for every participant. Participant IDs with an I prefix correspond to those recruited for the intervention part of the trial, while IDs with the C prefix indicate healthy controls. The dashed line indicates the binarisation threshold used for the daily symptom models. ML = Maximum likelihood; CV = Cross-validation.

One key assumption of this work is based on clinical evidence that the majority of ovulating individuals do not experience clinically significant shifts in affect due to their menstrual cycle^15^. We expect a similar effect for smartphone typing dynamics, in that the cycle-related fluctuations in BiAffect metrics reported above will be present in only a subset of the participants. Concretely, this means that we force one of the GAMM components in the mixture models to be (almost) completely flat by fixing its smoothing parameter to a high value. This restriction greatly increases the stability of the solutions found across iterations of the randomised cross-validation procedure we describe below (see also Supplementary Figure 6).

The smoothing parameters of the other, non-flat GAMM components can be found by either 1) using the standard GAMM procedures in case there is just one other component, or 2) doing a grid search over plausible parameter values with group k-fold cross validation (CV), ensuring each participant is assigned to only one of the folds. In simulation, this improves the mixture fit (Figure 2B, right panel).

Another challenge, which applies to nearly all mixture models, consists in selecting an adequate number of components^12^. Popular methods to address this issue include within-sample log-likelihood criteria (e.g., Akaike Information Criterion), but these would require corrections for models fitted with penalised likelihoods such as GAMMs^38^. While these corrections are available, it is not clear how they apply in a mixture model context. We therefore used the out-of-sample log-likelihood, calculated from a randomised group CV procedure, to aid the selection of the number of components (Figure 2A). We combined this with a qualitative assessment of the stability of the smooths fitted during each CV iteration. Mixture models with up to three components were subjected to these procedures to keep computational complexity manageable.

For movement rate, the one- and two-component mixture models show substantial stability across iterations, where the smooths fitted for the one-component model are consistent with the baseline movement rate model discussed above (Figure 2D). The two-component model, for which we forced one component to be flat, exhibits a curve that is similar to the one-component curve but with a larger amplitude. The attenuated amplitude of the one-component curve suggests that it is a compromise between participants that have a stable movement rate across the cycle and those that do not. We will refer to the first group of participants as flat, and to the second group as wiggly. The three-component model exhibits curves that are similar to those in the two-component model, with the addition of a set of curves with roughly three peaks: two in the luteal phase before menses, one in the follicular phase. Even though the out-of-sample log-likelihood favours the three-component model (Figure 2C), the variability of these curves is high, so we opted to continue with the two-component model.

Next, we calculated the responsibilities for the out-of-sample participants of all randomised CV iterations (Figure 2E). These responsibilities exhibited different behaviours for different participants, with some showing clear clustering and others showing frequent switching between extreme probability values. To facilitate downstream modelling, we took each participant’s average and binarised their responsibilities, such that those with an average component-2 responsibility greater than or equal to 0.5 were assigned to the wiggly group. These movement rate group assignments were then carried over to the daily rating analysis.

### Daily symptoms in typing subgroups

Finally, to relate the subtrajectories found in smartphone typing dynamics to clinical variation across the cycle, we created GAMMs for daily ratings of depression (Daily Record of Severity of Problems; DRSP)^39^, anxiety (DRSP), anhedonia (DRSP), suicidal ideation (a compound of several items from the Adult Suicidal Ideation Questionnaire (ASIǪ) and miscellaneous items)^40^, and irritability (DRSP). See the Supplement for a description of all items. The daily rating models required symptom and BiAffect data to align, which resulted in observations being dropped due to missing data. This meant that they were constructed from the data of 59 participants (49 with past-month SI, 10 controls) for depression, anxiety, anhedonia, and SI, and 58 participants (48 with past-month SI, 10 controls) for irritability. There were 3329 observations for depression (average: 56.4 per participant, range: 16-122), 3327 observations for anxiety, anhedonia, and SI (average: 56.4 per participant, range: 16-122), and 3293 for irritability (average: 56.8 per participant, range: 16-122).

The models consisted of a main effect of group assignment (i.e., intercept difference between flat and wiggly participants), a main effect of the cycle for the flat participants (cyclic smooth), a cycle-by-group interaction effect for the wiggly participants (difference smooth relative to the main cycle effect), and participant-level random intercepts and slopes. P-values of the fixed effects were adjusted for an FDR of 0.05. We used either Gaussian distributions with a log link (anxiety) or gamma distributions with an inverse link (depression, anhedonia, SI, irritability) as response distributions, depending on which resulted in the most Gaussian deviance residual distribution.

Figure 3 shows participant-centred averages of the daily ratings and the estimated GAMM responses for both groups. For the flat group, anhedonia and depression did not fluctuate significantly across the cycle (main effect smooth; anhedonia: edf = 0.00, F = 0.00, p = 0.95, p_adj_ = 0.96; depression: edf = 1.61, F = 1.34, p = 0.13, p_adj_ = 0.29), but these symptoms did show significant cycle-related fluctuations in the wiggly group (difference smooth; anhedonia: edf = 4.73, F = 8.65, p < 0.0001, p_adj_ = 0.00047; depression: edf = 4.73, F = 8.65, p < 0.0001, p_adj_ = 0.00047). Combined with the summed effects as displayed in Figure 3, this suggests that being part of the wiggly movement rate group is associated with larger deflections in anhedonia and depression ratings around menses. Anxiety, irritability, and SI, on the other hand, fluctuated significantly across the cycle for the flat group (main effect smooth; anxiety: edf = 3.34, F = 13.28, p = 0.00013, p_adj_ = 0.00047; SI: edf = 4.29, F = 9.12, p = 0.00017, p_adj_ = 0.00051; irritability: edf = 3.38, F = 4.45, p = 0.00011, p_adj_ = 0.00047) and did not show any significant deviations from those trajectories for the wiggly group (difference smooth; anxiety: edf = 0.00, F = 0.00, p = 0.96, p_adj_ = 0.96; SI: edf = 0.94, F = 0.31, p = 0.17, p_adj_ = 0.30; irritability: edf = 1.17, F = 0.28, p = 0.22, p_adj_ = 0.30), which implies similar curves for both participant groups. The main effect of group assignment was not significant for any rating (intercept difference; depression: β = 0.10, t(1) = 1.64, p = 0.10, p_adj_ = 0.25; anxiety: β = -0.11, t(1) = -0.97), p = 0.33, p_adj_ = 0.39; anhedonia: β = 0.088, t(1) = 1.29, p = 0.20, p_adj_ = 0.30; SI: β = 0.062, t(1) = 1.22, p = 0.22, p_adj_ = 0.30; irritability: β = 0.052, t(1) = 0.96, p = 0.34, p_adj_ = 0.39).

**Figure 3:**
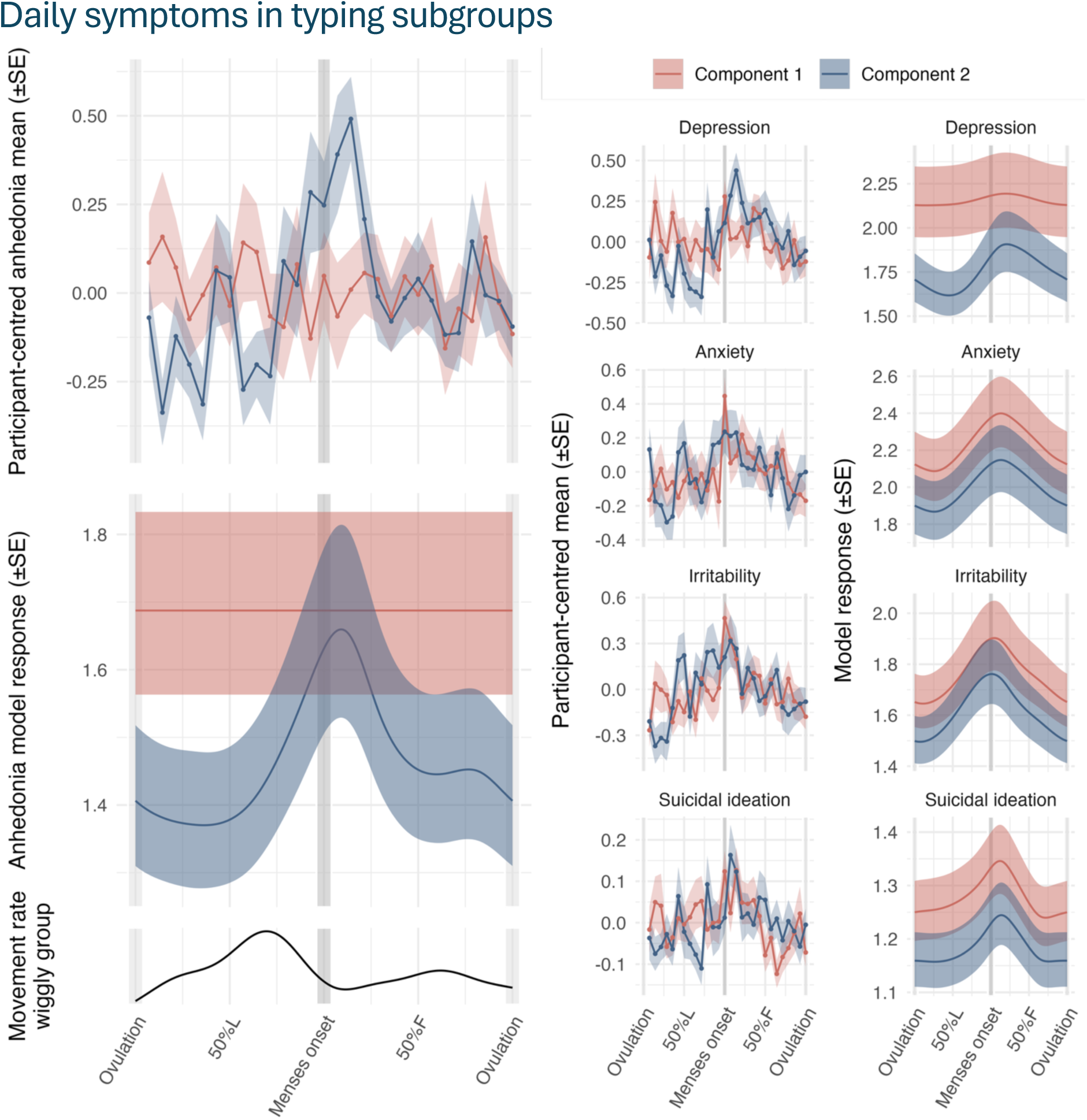
Participant-centred symptom means and mean responses estimated by GAMMs across the menstrual cycle. Top- and middle-left panels: Participant-centred means and associated GAMM responses for anhedonia ratings, moderated by the latent trajectory grouping found by the smartphone movement rate mixture models. Bottom-left panel: Mean movement rate response of the wiggly group component models (Figure 2D, middle panel, blue lines), added to allow comparison of temporal developments in smartphone movement and daily ratings. Right panels: Participant-centred means and model responses for depression, anxiety, irritability, and suicidal ideation.

All model residuals showed slight deviations from normality, which we could not improve with alternative response distributions or link functions; this might be partly due to the skewed, discrete distribution of the Likert scale responses. Deviations were more pronounced for the anhedonia model and rather severe for the SI model (see Supplementary Figure 7).

## Discussion

In this work, we set out to parse inter-individual differences in digital phenotyping measures of behaviour and relate these to meaningful differences in psychiatric symptom dynamics. We demonstrate this approach in the context of non-linear affective change over the menstrual cycle using a transdiagnostic sample of healthy controls and outpatients with past-month suicidal ideation, where participants’ smartphone typing dynamics were passively recorded for the duration of the study along with daily symptom ratings. After first characterising baseline typing fluctuations across the cycle for the entire sample, we found stable latent trajectories in smartphone movement rate while typing with a novel latent trajectory mixture modelling approach: one group of participants showed minimal behavioural fluctuation over the cycle, whilst the other showed amplified movement rate fluctuations compared to the baseline model. Those participants with amplified movement fluctuations also had increased fluctuations in symptoms across the menstrual cycle, specifically in anhedonia and depression ratings, but not in anxiety, irritability, and suicidal ideation (SI).

Parsing inter-individual variation is at the core of our approach. This is in line with the precision medicine initiative, which aims to use individual variability to inform diagnosis and, ultimately, treatment selection^41^. In the context of digital phenotyping, this requires identifying the variability in the digital phenotype that is relevant to psychopathology and separating that from other sources of variability, which we achieved with a growth mixture modelling method. This yielded several common patterns of typing dynamics across the menstrual cycle as well as the degree to which each participant was associated with each pattern. In order for such a stratification to be useful, however, it is necessary to demonstrate that it relates to clinically meaningful variation. We therefore showed that these latent typing trajectories map differentially onto the development of symptoms across the menstrual cycle. Taken together, this approach is a first step towards a precision medicine approach for digital phenotyping in menstrual cycle research. The passive, unobtrusive, and highly cost-effective nature of smartphone-based digital phenotyping make it attractive to explore further as a supporting diagnostic tool that might alleviate the burden of daily symptom ratings.

The results found in this study align broadly with emerging frameworks highlighting individual differences in sensitivity to the cycle. For instance, the Dimensional Affective Sensitivity to Hormones across the Menstrual Cycle (DASH-MC) Framework draws on experimental evidence to posit differences in affective reactivity to both oestrogen (E2) and progesterone (P4) change across the cycle^42^. These differences in cyclical negative affect are hypothesized to stem from a combination of heightened sensitivity to progesterone metabolite surges during the luteal phase (commonly but not exclusively associated with irritability and other high-arousal affect) and oestradiol withdrawal sensitivity at menstruation (linked to depressive/anhedonic mood, cognitive impairments, and fatigue). Within the present study, the phenotypic findings linking reduced movement around times of oestradiol withdrawal (immediately after ovulation and during menstruation) to increased depressive affect are suggestive of a role for acute oestradiol fluctuation or withdrawal. Conversely, the increase in movement across the luteal phase raises the possibility of an additional mechanism related to progesterone metabolite flux. Ultimately, a larger sample size combined with experimental hormone manipulations would be necessary to distinguish between these unique pathways. At present, it is most prudent to hypothesise a greater affective sensitivity to hormones among those exhibiting cyclical movement flux.

Whilst this study ultimately cannot speak to the mechanisms underlying the potential link between movement and depressive flux, future work could utilise these passive sensing approaches to investigate whether their ability to passively index affective change is mediated by shared biological systems. To illustrate this point and stimulate further hypothesis generation, we provide a speculative interpretation by considering the construct of psychomotor retardation, a well-documented feature of depression^43–45^. Psychomotor retardation in depression has often been hypothesised to relate to dopaminergic deficits, with studies showing impaired pre- and postsynaptic striatal dopamine function specifically for psychomotor retardation in depression^46–49^. Dopamine also plays an important role in how the body responds to changes in oestrogen levels. Animal studies have shown, for instance, that both natural and induced increases in oestrogen lead to increased dopamine availability and striatal receptor density^42,50–52^. To explore the interplay between these components, future studies could use digital phenotyping to assess additional measures of psychomotor retardation while incorporating repeated urinary oestrogen (E1G) measures as well as laboratory measures of dopaminergic function.

There are several limitations to our study. First, all multicomponent mixture models featured a component which we forced to fit a (nearly) flat line. While this operationalised our prior assumption that a portion of the participants would not show meaningful fluctuations in typing metrics across the cycle, one could argue that a more elegant solution would be to put no flatness constraints on the components and find the latent trajectories in a fully data-driven fashion. Unfortunately, such an approach has to overcome several practical obstacles. Not constraining one of the components increases the (inner) k-fold CV parameter grid with an additional dimension, quickly leading to prohibitively large computational costs for the (outer) randomised CV. Moreover, the ordering of components within a mixture model is arbitrary, which makes it difficult to line them up when examining the stability of the trajectories or averaging the responsibilities (Supplementary Figure 6).

Second, as with all mixture models, selecting an appropriate number of components remains challenging. Whilst the median out-of-sample log-likelihood of the three-component movement rate solution was highest, its latent trajectories appeared decidedly more unstable in our visual assessment. The two-component model seemed like a suitable compromise between these two factors. Note, however, that we do not claim that this number is optimal to describe the entire population, nor that these components necessarily have a direct biological interpretation. The instability of the three-component model might have come about because of our limited sample size, and larger sample sizes could potentially provide evidence for more components.

Finally, it should be noted that this study was highly explorative and intended for generating hypotheses and developing new research tools. More research in larger samples is needed to determine the relevance of the latent movement rate trajectories, whether the relationship with depression and anhedonia holds up in larger samples, and if so, the causal mechanism behind this relationship. Furthermore, the effects we discovered are group effects, and additional work needs to be done to ascertain if smartphone movement while typing is a useful metric to estimate dimensions of affective hormonal sensitivity in an individual.

In summary and with these limitations notwithstanding, we have provided a methodology for differentiating clinical trajectories by parsing inter-individual heterogeneity in unobtrusively collected digital phenotyping data. This procedure is not specific to the measures analysed here and offers the modeller a useful tool to understand the patterns underlying digital phenotyping data collected from heterogeneous samples.

## Methods

### Study design

The CLEAR-3 trial was a randomised clinical crossover trial that investigated the hormonal mechanisms behind perimenstrual worsening of suicidal ideation and affective symptoms (NCT04112368). The effects of natural steroid withdrawal under placebo were compared to those of hormonal flux stabilisation by oestradiol (E2) and progesterone (P4) administration. After a baseline period of at least one full menstrual cycle, each intervention participant was randomised to either two weeks of placebo or two weeks of E2+P4 administration. Participants then entered a wash-out period of one menstrual cycle before crossing over to the other condition. In this study, we only used data from the baseline and wash-out periods. The study was approved by the UIC Institutional Review Board and all relevant ethical regulations were adhered to.

### Recruitment and exclusion criteria

All participants provided informed consent for study participation. They were recruited from the community using social media ads and received $1250 upon study completion. Inclusion criteria required participants to be 18-45 years of age; have menstrual cycles between 25-35 days; and have a BMI between 18-29. Participants in the intervention group were required to have reported at least some suicidal ideation in the past month at the time of recruitment while also having an acceptably low imminent risk for a suicide attempt. They additionally had to be under the current care of an outpatient mental health provider. Exclusion criteria included breastfeeding, being pregnant, or trying to become pregnant; using any exogenous or intrauterine hormonal medications; any long-term non-psychiatric health condition; cigarette smoking; any history of manic episodes or psychotic symptoms; a current substance use disorder; a history or diagnosis of postpartum depression; and a history or diagnosis of premenstrual dysphoric disorder (as evidence-based treatments are available).

### Data preprocessing

BiAffect data were preprocessed as in previous works^23,27,28^. Individual key presses were first aggregated into typing sessions, which are initiated when a user opens the keyboard and terminated when the keyboard closes or after six seconds of inactivity. We calculated session-level summary metrics of the inter-key delays (IKDs), i.e., median IKD, median absolute deviation (MAD) IKD, and the 95^th^ percentile IKD.

We additionally used the phone’s triaxial accelerometer data to determine the orientation of the phone (upright rate) and whether it was moving (movement rate). These were only collected when the keyboard was active at a sampling rate of 10Hz. High-frequency noise was filtered out of the signal using a second-order bidirectional Butterworth filter with a 4 Hz cut-off. An entire typing session was classified as stationary if at least 92% of all accelerometer samples had a magnitude within 0.95 and 1.05 (inclusive bounds), where a magnitude of 1 represents the pull of the earth; otherwise, the session was classified as moving. Furthermore, sessions were classified as upright if the median accelerometer value in the x-direction was between -0.2 and 0.2 (inclusive bounds) and the median value in the z-direction was lower than 0.1.

Session-level variables were then aggregated into daily variables to match the temporal resolution of the psychopathology ratings. Filtering requirements that determined which typing sessions were included into the aggregation differed from variable to variable:

1. The total number of key presses (including autocorrect events): no filtering. Aggregation took place by summing the session-level number of key presses.
2. Movement and upright rate: sessions were required to have at least twenty key presses. Session-level moving and upright classifications were counted and then divided by the total number of sessions recorded for that day to get movement and upright rate, respectively.
3. All IKD measures, autocorrect rate, and backspace rate: sessions had to have at least twenty key presses, be typed with two hands, and be upright and stationary. Daily-level IKD measures were calculated by taking the mean of the corresponding session-level measures. Autocorrect events and backspace presses were counted and divided by the total number of key presses found in all eligible typing sessions to give autocorrect and backspace rates. In addition, typing speed instability was calculated as the root-mean-square of successive differences (RMSSD) of the session-level median IKD values.

### Generalised additive mixed models

Generalised additive mixed models (GAMMs) are generalised linear mixed models (GLMMs; not to be confused with *general* linear models) with a linear predictor that contains a sum of quadratically penalised smooth functions of covariates^31,32^. This means that, just like any other GLMM, they use an exponential family response distribution along with a link function *g*:

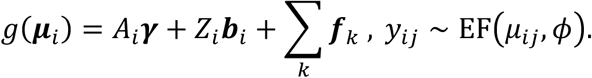

Here, the *j*th response of participant *i*, *y_i_*_j_, is assumed to follow an exponential family distribution with mean *μ_i_*_j_ and scale parameter *φ*. A participant’s *μ_i_*_j_ are collected in a (column) vector ***μ****_i_* = [*μ_i_*_1_, … , *μ_ini_*]^T^ of length *n* and are modelled by participant *i*’s parametric model matrix *A_i_*; parametric, fixed-effects coefficient vector ***γ***; random effect model matrix *Z_i_*; and random effect coefficients ***b****_i_*. Finally, a sum of evaluated smooth functions is added, each of which is calculated as:

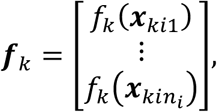

where ***x****_ki_*_j_ is a scalar or vector of covariates (that often coincides with part of the rows of *A_i_*, but this is not necessary).

More concretely, for the baseline typing dynamics models, the regression function was as follows:

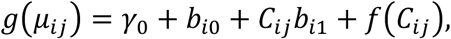

where *γ*_0_ and *b_i_*_0_ are, respectively, the fixed and random intercept, and *b_i_*_1_ is the random slope of the PACTS cycle estimate *C_i_*_j_ ∈ (−1, 1]. *f* is a cyclic cubic regression spline, configured such that -1 and 1 are considered the same point in covariate space. The PACTS cycle estimates were calculated with the menstrualcycleR package (version 0.1.2).

In general, the smooth functions are often expressed as overparameterised basis expansions, where the quadratic penalisation prevents overfitting. The amount of penalisation is governed by a smoothing parameter, which is found using techniques such as generalised cross-validation (GCV), maximum likelihood (ML), or restricted maximum likelihood (REML)^32^; for the baseline models, we chose to use REML, as it is less prone to undersmoothing than, for instance, GCV^32^.

For null hypothesis testing, it should be noted that smooth p-values are approximate and do not take the uncertainty in smoothing parameter estimates into account^53^. Correction for multiple comparisons was performed with the Benjamini-Hochberg procedure^54^.

Outliers were only excluded if their associated deviance residuals deviated strongly from a normal distribution (assessed with Q-Q plots) and if the total number of deviating residuals was lower than five. This was the case only for autocorrect and backspace rate models. Removing the associated data points did not substantially alter the model fit.

### Mixture models

Mixture models are typically fitted using expectation maximisation (EM), which is an iterated two-step procedure to find maximum likelihood solutions for models with latent variables^37^. In the case of mixture models, the latent variable *H* specifies which component every observation or group of observations is assigned to. Here, *H* represents a collection of one-hot-encoded vectors ***h****_i_*, which are vectors that have a value of one at exactly one location and zeros everywhere else. The location of the value one specifies which component participant *i* belongs to. For instance, if a participant belongs to the second component in a three-component mixture model, then ***h****_i_* = [0 1 0].

The E step of the EM algorithm consists of re-estimating which assignments are most likely given the data X, ***y*** and all component model parameters *θ*^old^ by evaluating *p*(*H* ∣ X, ***y***, *θ*^old^). In the context of GAMMs, X refers to the collection of all model matrices (*A_i_*, *Z_i_*, and the vectors ***x****_k_*_j*i*_) across all participants and *θ*^old^ denotes all parameters (***γ***, ***b****_i_*, the parameters involved in the basis expansion of the *f_k_*, and potentially the smoothing parameter). This probability can be calculated for one specific component *m* and participant *i* as 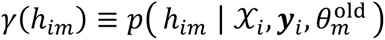, in which X*_i_*, ***y****_i_* are those subsets of X, ***y*** that correspond to participant *i*, ℎ*_im_* is the *m*th entry of ***h****_i_*, and 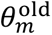 is the subset of *θ*^old^ corresponding to the *m*th component model. *γ*(ℎ*_im_*) is also known as the *responsibility* that component *m* takes for the data of participant *i*.

The M step then consists of finding new model parameters *θ*^new^ by optimising the component models based on the updated responsibilities. This can be done by finding the expectation of the joint component model log likelihood w.r.t. the responsibilities (posterior of the component assignments) calculated in the E step. More specifically,

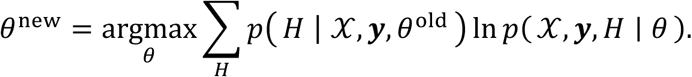

We use Σ*_H_* as a shorthand for Σ***_h_***_1∈ℋ_ … Σ***_h_****_N_*_∈ℋ_, where *N* is the number of participants and ℋ is the set of all possible one-hot vectors that the ***h****_i_* could take as their value. In other words, the expectation is calculated using all possible participant-component assignments.

The E and M steps are then repeated until convergence. Mixture models were fitted using the R flexmix package (version 2.3-20). We adapted the source code of the FLXMRmgcv class to allow fixing the smoothing parameters to prespecified values, which was necessary for inducing flatness (by fixing the parameter to a high value) and k-fold cross-validation (see below). In other cases, smoothing parameters were fitted using the default method in flexmix, namely maximum likelihood.

flexmix only allows response distributions that are Gaussian, binomial, and Poisson, which means that median IKD and log RMSSD median IKD had to be modelled with a Gaussian distribution rather than a scaled t distribution (which may have been preferable), and movement rate and autocorrect were modelled with binomial distributions instead of quasibinomial ones.

### k-fold cross-validation

As discussed above, GAMMs find their smoothing parameters *λ* through, for instance, (RE)ML or GCV. The theory that these procedures are based on, however, does not specify how to find the smoothing parameters across multiple GAMMs in a mixture context. This was necessary in our three-component mixture models: Although these models had the smoothing parameter of one component fixed to a high value (inducing flatness), the smoothing parameters of the remaining two components still required simultaneous estimation. We opted to solve this issue computationally, with five-fold cross-validation (CV) over a grid of log *λ* values, ensuring participants were assigned to exactly one fold. Every log *λ* that required fitting had five candidate values (-7, -3.5, 0, 3.5, and 7) and the grid consisted of all possible combinations of those values. While five values per log *λ* is coarse, it allowed us to keep computational complexity manageable. The solution with minimal average negative log-likelihood across all the testing folds was then fitted to all data under consideration (i.e., all folds).

Calculating the negative log-likelihood out of sample (that is, on the test fold) requires integrating out the latent component assignment *H* while taking the data’s group structure (observations within participants) into account. Note that we set random effects to zero during these calculations. To evaluate the log-likelihood, we start with the density of the test data X^∗^, ***y***^∗^, given the parameters of the component models:

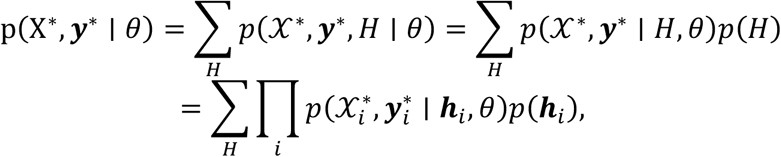

where 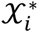 and 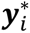 are, respectively, the subset of the model matrices and response vector corresponding to test participant *i*. The one-hot structure of ***h****_i_* effectively allows us to select which component model *m* to use when calculating the log-likelihood:

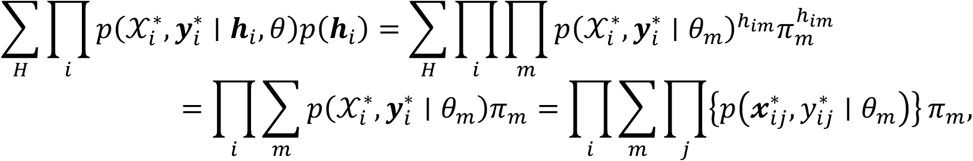

where ℎ*_im_* ∈ {0, 1} is the *m*th element of ***h****_i_*, *π_m_* is the prior probability of data belonging to component *m*, and *θ_m_* are the parameters associated with the model of component 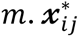 is the vector containing all test data for the *j*th observation of participant *i*, and 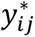 is the associated response. This quantity is easily calculated from the fitted component models, but we take its log to prevent numerical underflow:

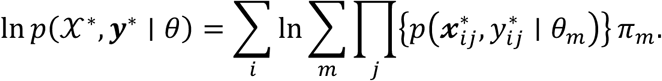

The sum of products inside the log can be evaluated safely using the log-sum-exp function.

### Randomised cross-validation

We used randomised CV to assess the stability of the mixture model solutions and select an appropriate number of mixture components. On every iteration, roughly 30% of all participants are sampled into the test set. One-, two-, and three-component mixture models are then fitted on the remaining 70% of participants. The component model equations are the same as for the baseline models described above. For the one-component model, the smoothing parameter of *f* was found through ML. For the two-component model, the smoothing parameter of one component was fixed to a high value (to induce flatness), whilst the other was still found through ML. For the three-component model, one parameter was fixed to a high value as for the two-component model but the smoothing parameters for the remaining two components were found through 5-fold CV as described above. The randomised CV procedure was run with 1000 iterations.

### Daily symptom models

The daily symptom models are similar to the baseline GAMMs used for smartphone typing dynamics, except that they now additionally feature a moderation by the movement rate groups discovered through mixture modelling:

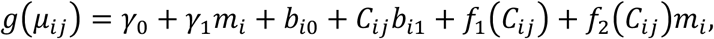

where *m_i_* is a binary variable indicating movement rate latent trajectory membership; *m_i_* = 1 for participants with movement rate fluctuations (the “wiggly” group), and *m_i_* = 0 for those without (the “flat” group). *γ*_1_ is the intercept offset associated with the wiggly group. *f*_1_ is the main effect of the cycle on the symptom being modelled, and *f*_2_ is the difference from that effect associated with the wiggly group (referred to as the difference smooth in the main text).

### Simulation

To demonstrate that using the default procedures to find GAMM smoothing parameters can be suboptimal in a mixture modelling context and the consequent value of our k-fold CV approach, we used both procedures to fit mixture models to data simulated from two clusters. The data were constructed with random intercepts and slopes and clusters consisted of either a straight line or a sine wave:

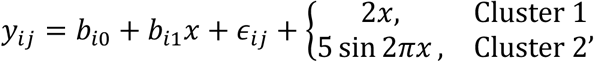

where *x* ∼ *U*(−0.5, 0.5), *b_i_*_0_ ∼ N(0, 1^2^), *b_i_*_1_ ∼ N(0, 2^2^), and *ε_i_*_j_ ∼ N(0, 5^2^). Data were simulated for 20 groups (which can be interpreted as participants), each with 28 observations. The probabilities for participants to be sampled into Clusters 1 and 2 were 0.4 and 0.6, respectively.

We then fitted the data with the two different approaches. All component models used the following model equation:

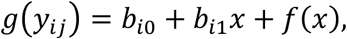

where *g* was the identity link and *y_i_*_j_ was assumed to be normally distributed. *f* was represented by a thin-plate spline. The default approach used ML to find the smoothing parameters *λ*. The CV approach used five folds and the same log *λ* grid as the BiAffect data models.

## Data availability

Deidentified participant data will be made available on reasonable request to the principal investigator of the CLEAR-3 trial, T.A. Eisenlohr-Moul.

## Code availability

All analysis source code is freely available on GitHub: https://github.com/Valkje/clear3-subtrajectories.

## Acknowledgements

This study was funded by the European Research Council (101001118). TAE-M, AL, and AN received funding from the National Institute of Mental Health (RF1MH120843, R01MH120168, and F30MH138058, respectively). CFB gratefully acknowledges funding from the Wellcome Trust Collaborative Award in Science 215573/Z/19/Z and the Netherlands Organisation for Scientific Research Vici Grant No. 17854 and NWO-CAS Grant No. 012-200-013.

## Author contributions

LK: Conceptualisation; data curation; software; formal analysis; methodology; visualisation; writing – original draft. AN: Data curation; methodology; writing – review & editing. FH: Software. CFB: Funding acquisition; supervision. AL: Conceptualisation; funding acquisition; investigation; methodology; supervision; writing – review & editing. TAE-M: Conceptualisation; data curation; funding acquisition; methodology; investigation; writing – review & editing. AFM: Conceptualisation; funding acquisition; methodology; supervision; writing – review & editing.

