## Supplement for "Parsing inter-individual variability in the digital phenotype across the menstrual cycle"

### Demographics

***Supplementary Table 1***

*Demographics of the various samples used in analysis. Baseline and mixture sample characteristics were calculated for the movement rate models. Symptom sample characteristics were calculated for the anhedonia model.*

| | Baseline intervention (N = 56) | Baseline controls (N = 13) | Mixture intervention (N = 52) | Mixture controls (N = 10) | Symptom intervention (N = 49) | Symptom controls (N = 10) |
|---|---|---|---|---|---|---|
| **Age (mean (SD))** | 27.57 (4.67) | 26.73 (5.61) | 27.75 (4.69) | 26.12 (4.32) | 27.61 (4.77) | 26.12 (4.32) |
| **Ethnicity (N (%))** | | | | | | |
| Hispanic | 14 (0.25) | 2 (0.15) | 13 (0.25) | 2 (0.2) | 11 (0.22) | 2 (0.2) |
| Non-Hispanic | 40 (0.71) | 7 (0.54) | 38 (0.73) | 4 (0.4) | 37 (0.76) | 4 (0.4) |
| Unknown or not reported | 2 (0.04) | 4 (0.31) | 1 (0.02) | 4 (0.4) | 1 (0.02) | 4 (0.4) |
| **Gender (N (%))** | | | | | | |
| Female | 51 (0.91) | 9 (0.69) | 48 (0.92) | 6 (0.6) | 45 (0.92) | 6 (0.6) |
| Nonbinary | 4 (0.07) | 0 (0.00) | 3 (0.06) | 0 (0.00) | 3 (0.06) | 0 (0.00) |
| Other | 1 (0.02) | 0 (0.00) | 1 (0.02) | 0 (0.00) | 1 (0.02) | 0 (0.00) |
| Unknown or not reported | 0 (0.00) | 4 (0.31) | 0 (0.00) | 4 (0.4) | 0 (0.00) | 4 (0.4) |
| **Household income (N (%))** | | | | | | |
| less than $15,000 | 4 (0.07) | 0 (0.00) | 4 (0.08) | 0 (0.00) | 4 (0.08) | 0 (0.00) |
| $15,000 - $19,999 | 1 (0.02) | 0 (0.00) | 1 (0.02) | 0 (0.00) | 1 (0.02) | 0 (0.00) |
| $20,000 - $24,999 | 3 (0.05) | 0 (0.00) | 3 (0.06) | 0 (0.00) | 3 (0.06) | 0 (0.00) |
| $25,000 - $29,999 | 3 (0.05) | 0 (0.00) | 3 (0.06) | 0 (0.00) | 3 (0.06) | 0 (0.00) |
| $30,000 - $34,999 | 2 (0.04) | 0 (0.00) | 2 (0.04) | 0 (0.00) | 2 (0.04) | 0 (0.00) |
| $35,000 - $39,999 | 3 (0.05) | 1 (0.08) | 3 (0.06) | 1 (0.1) | 3 (0.06) | 1 (0.1) |
| $40,000 - $49,999 | 3 (0.05) | 0 (0.00) | 3 (0.06) | 0 (0.00) | 3 (0.06) | 0 (0.00) |

| | | | | | | |
|---|---|---|---|---|---|---|
| $50,000 - $79,999 | 16 (0.29) | 4 (0.31) | 14 (0.27) | 2 (0.2) | 12 (0.24) | 2 (0.2) |
| $80,000 - $99,999 | 7 (0.12) | 1 (0.08) | 6 (0.12) | 0 (0.00) | 6 (0.12) | 0 (0.00) |
| $100,000 or above | 8 (0.14) | 2 (0.15) | 8 (0.15) | 2 (0.2) | 8 (0.16) | 2 (0.2) |
| Unknown or not reported | 6 (0.11) | 5 (0.38) | 5 (0.1) | 5 (0.5) | 4 (0.08) | 5 (0.5) |
| **Race (N (%))** | | | | | | |
| African American | 7 (0.12) | 0 (0.00) | 7 (0.13) | 0 (0.00) | 6 (0.12) | 0 (0.00) |
| Asian | 6 (0.11) | 6 (0.46) | 5 (0.1) | 3 (0.3) | 5 (0.1) | 3 (0.3) |
| Caucasian | 28 (0.5) | 2 (0.15) | 26 (0.5) | 2 (0.2) | 26 (0.53) | 2 (0.2) |
| More than one race | 6 (0.11) | 0 (0.00) | 6 (0.12) | 0 (0.00) | 6 (0.12) | 0 (0.00) |
| Unknown or not reported | 9 (0.16) | 5 (0.38) | 8 (0.15) | 5 (0.5) | 6 (0.12) | 5 (0.5) |
| **Baseline Diagnosis (N (%))** | | | | | | |
| Any current anxiety disorder | 31 (0.55) | 0 (0) | 30 (0.58) | 0 (0) | 28 (0.57) | 0 (0) |
| Any current bipolar disorder | 2 (0.04) | 0 (0) | 2 (0.04) | 0 (0) | 2 (0.04) | 0 (0) |
| Current ADHD | 11 (0.2) | 0 (0) | 9 (0.17) | 0 (0) | 9 (0.18) | 0 (0) |
| Current borderline personality disorder | 4 (0.07) | 0 (0) | 4 (0.08) | 0 (0) | 4 (0.08) | 0 (0) |
| Any current depressive disorder | 35 (0.62) | 0 (0) | 31 (0.6) | 0 (0) | 28 (0.57) | 0 (0) |
| Any current eating disorder | 3 (0.05) | 0 (0) | 3 (0.06) | 0 (0) | 2 (0.04) | 0 (0) |
| Any current obsessive-compulsive disorder | 4 (0.07) | 0 (0) | 4 (0.08) | 0 (0) | 4 (0.08) | 0 (0) |
| Any current substance use disorder | 11 (0.2) | 0 (0) | 10 (0.19) | 0 (0) | 9 (0.18) | 0 (0) |
| Any current trauma-related disorder | 11 (0.2) | 0 (0) | 10 (0.19) | 0 (0) | 9 (0.18) | 0 (0) |

### Model sample sizes

***Supplementary Table 2***

*Sample sizes for the various baseline GAMMs and mixtures of GAMMs. A hyphen indicates no mixture model was fitted.*

| | Baseline sample size | Mixture sample size |
|---|---|---|
| Movement rate | 69 (13 controls), 3454 observations | 62 (10 controls), 3356 observations |
| Upright rate | 69 (13 controls), 3454 observations | 62 (10 controls), 3356 observations |
| Autocorrect rate | 65 (11 controls), 3170 observations | 60 (10 controls), 3107 observations |
| Backspace rate | 65 (11 controls), 3170 observations | 60 (10 controls), 3107 observations |
| Median IKD | 65 (11 controls), 3155 observations | 59 (9 controls), 3080 observations |
| MAD IKD | 65 (11 controls), 3155 observations | 59 (9 controls), 3080 observations |
| 95th percentile IKD | 65 (11 controls), 3170 observations | 60 (10 controls), 3107 observations |
| RMSSD IKD | 62 (10 controls), 2909 observations | 58 (9 controls), 2844 observations |
| # key presses | 72 (13 controls), 3574 observations | 65 (11 controls), 3478 observations |

### Baseline correlations and model estimates

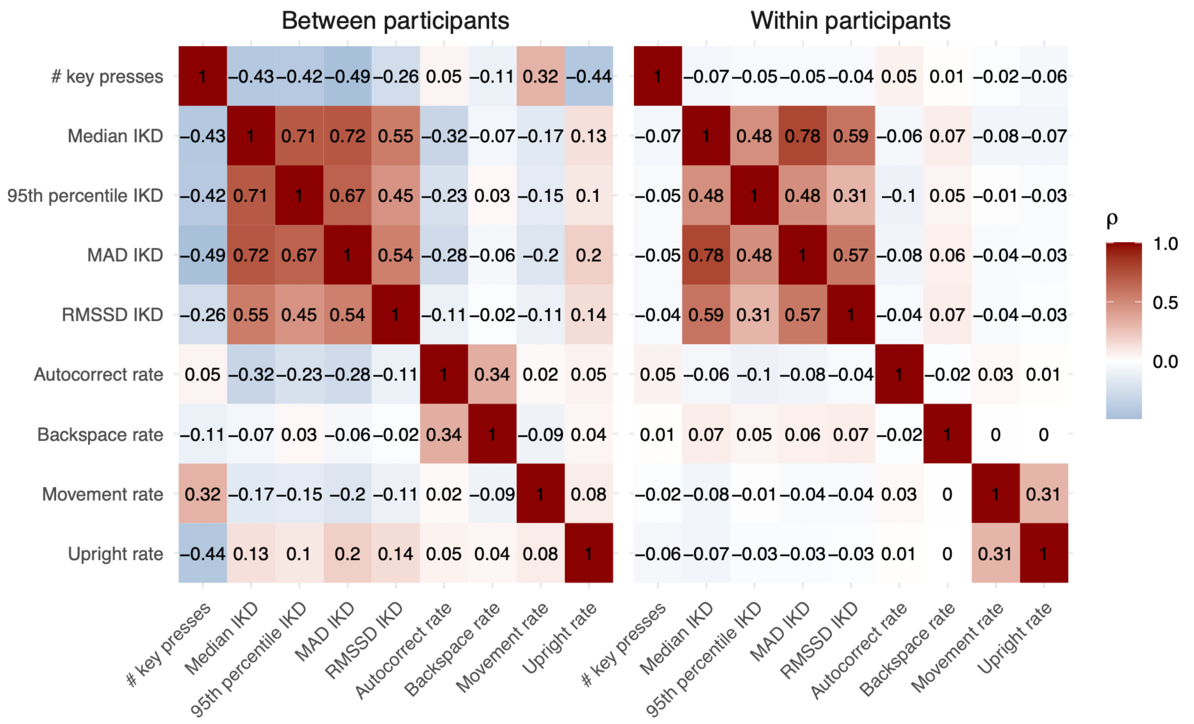

***Supplementary Figure 1: Between- and within-participant correlations of BiAffect measures.*** *Between-participant correlations are Spearman's rank correlations of participant means of every BiAffect measure. Within-participant correlations were calculated using the R package rmcorr (version 0.7.0). Participants were included if they had at least 5 days of complete BiAffect data (regardless of menstrual cycle phase), resulting in 9160 observations for 117 participants.*

***Supplementary Table 3***

*Model estimates and associated statistics of menses-centred baseline GAMMs. F is a weighted sum of chi-squared statistics. Benjamini-Hochberg adjustment was only applied to cycle effect p-values as those were the only effects of interest to us; (uncorrected) random effect p-values are reported for completeness. edf = Effective degrees of freedom.*

| Response | Cycle effect | Random intercept | Random cycle slope |
| --- | --- | --- | --- |
| Movement rate | edf = 4.13, F = 12.49, p = 0.00011, $p_{adj}$ = 0.0010 | edf = 62.57, F = 50.52, p < 0.0001 | edf = 23.84, F = 2.61, p = 0.00053 |
| Autocorrect rate | edf = 2.25, F = 3.82, p = 0.041, $p_{adj}$ = 0.092 | edf = 61.26, F = 71.18, p < 0.0001 | edf = 25.64, F = 7.53, p < 0.0001 |
| Backspace rate | edf = 0.0087, F = 0.00065, p = 0.57, $p_{adj}$ = 0.76 | edf = 61.39, F = 77.32, p < 0.0001 | edf = 23.43, F = 4.08, p = 0.015 |
| MAD IKD | edf = 0.0017, F = 0.00031, p = 0.88, $p_{adj}$ = 0.88 | edf = 63.34, F = 7852.58, p < 0.0001 | edf = 0.005, F = 0.0039, p = 0.88 |
| Median IKD | edf = 1.66, F = 20.49, p = 0.012, $p_{adj}$ = 0.053 | edf = 63.5, F = 11043.18, p < 0.0001 | edf = 12.01, F = 67.36, p = 0.023 |
| 95th percentile IKD | edf = 0.00028, F = 0.00016, p = 0.59, $p_{adj}$ = 0.76 | edf = 61.81, F = 2577.25, p < 0.0001 | edf = 23.8, F = 83.79, p = 0.0027 |
| Log RMSSD IKD | edf = 1.71, F = 5.44, p = 0.035, $p_{adj}$ = 0.092 | edf = 53.24, F = 502.04, p < 0.0001 | edf = 0.018, F = 0.016, p = 0.65 |
| Total number of key presses | edf = 0.061, F = 0.006, p = 0.48, $p_{adj}$ = 0.76 | edf = 69.92, F = 65.58, p < 0.0001 | edf = 31.43, F = 3.17, p = 0.021 |
| Upright rate | edf = 0.00093, F = 5.1e-05, p = 0.68, $p_{adj}$ = 0.76 | edf = 61.36, F = 41.87, p < 0.0001 | edf = 17.92, F = 1.16, p = 0.059 |

### Mixture models for other BiAffect metrics

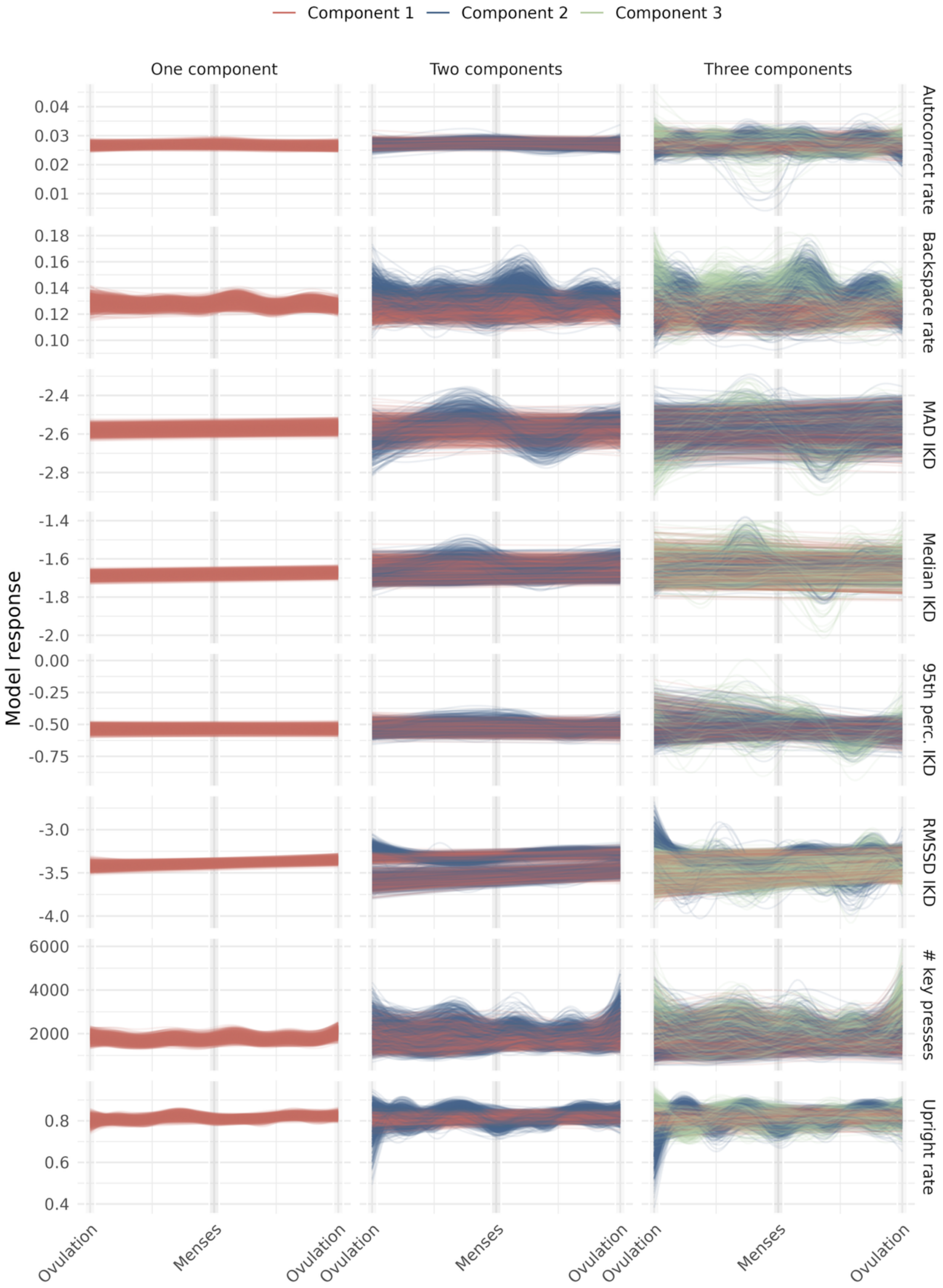

***Supplementary Figure 2: Stability analysis results for mixture models of eight BiAffect typing metrics.*** *Rows correspond to a BiAffect metric, and columns give results for a one-, two-, or three-component mixture model. Every line plotted in these figures gives the model fit from one iteration of the stability analysis algorithm described in the main text. All IKD metrics were log-transformed before the mixture models were fitted, hence their negative model responses.*

We ran our stability analysis on one-, two-, and three-component mixture models of eight other BiAffect metrics (Supplementary Figure 2). Out-of-sample log-likelihoods (not shown) indicated that higher-order mixture models were preferred for the total number of key presses, upright rate, backspace rate, and autocorrect rate. Of these four, only the two-component models for upright rate and backspace rate showed both stable component curves (Supplementary Figure 2) and somewhat stable component responsibilities (not shown).

We therefore tested if the two-component responsibilities generated by the upright and backspace rate mixture models would be predictive of depressive symptom trajectories (Supplementary Figure 3), as we did with the movement rate responsibilities in the main text. GAMMs indicated that, when compared to individuals assigned to component 1 of the mixture models, individuals assigned to component 2 displayed a slightly different depression severity trajectory for backspace rate (edf = 2.83, F = 4.21, p = 0.016), but not for upright rate (edf = 0.00, F = 0.00, p = 0.71). We additionally tested if the significant backspace rate moderation would extend to anhedonia ratings, but this was not the case (edf = 0.68, F = 0.11, p = 0.38).

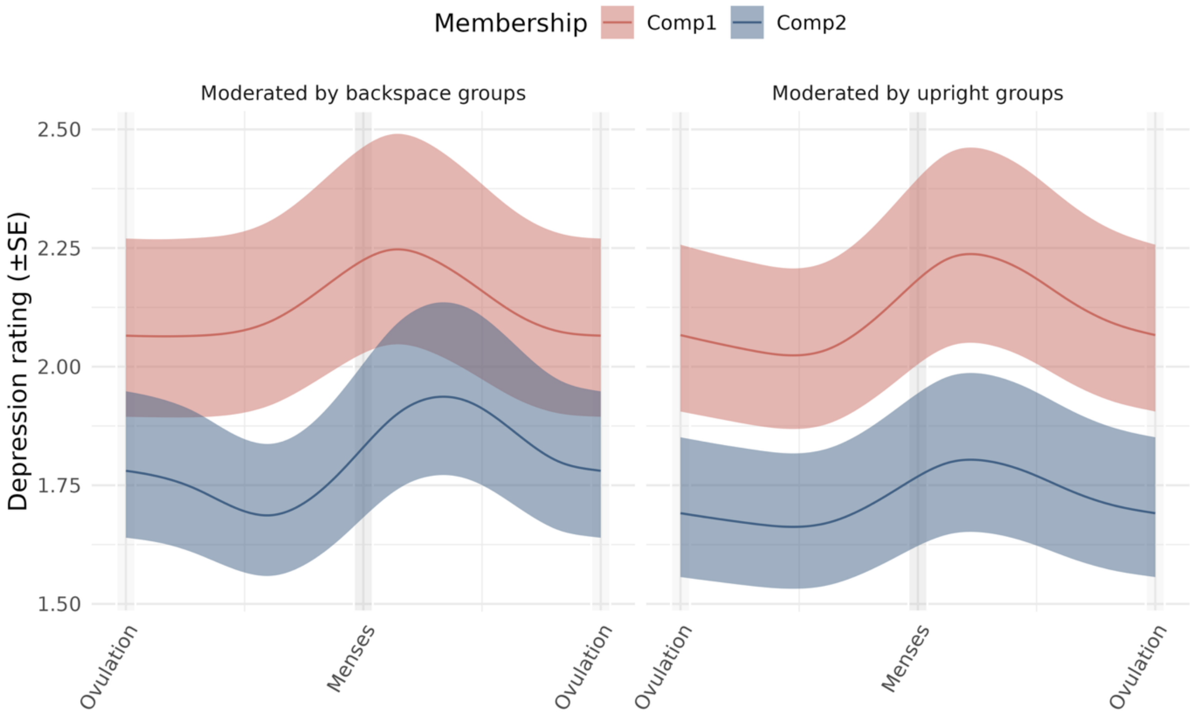

***Supplementary Figure 3: Menstrual cycle effects on depression, moderated by groups defined by the backspace and upright rate mixture models.*** *The left panel, which gives the backspace rate group moderation, shows the difference in depression trajectories for the two groups. This effect is not present in the right panel, which shows the upright rate group moderation.*

The interpretation of the backspace rate group moderation on depression across the cycle is more challenging than for movement rate. The curves for the second component of the two-component backspace rate model (Supplementary Figure 2) show high wiggliness, with increases midluteally, postmenstrually, and preovulatory. This much variability, which was not apparent in the backspace rate base model, could be due to poor model fit. Moreover, the difference between the depression rating trajectories seems to be only slight (Supplementary Figure 3), with the component 2 group featuring a small premenstrual dip in depression rating before showing the same peri-

/postmenstrual increase as the component 1 group. The clinical relevance of this difference is unclear.

### Ovulation-centred baseline models

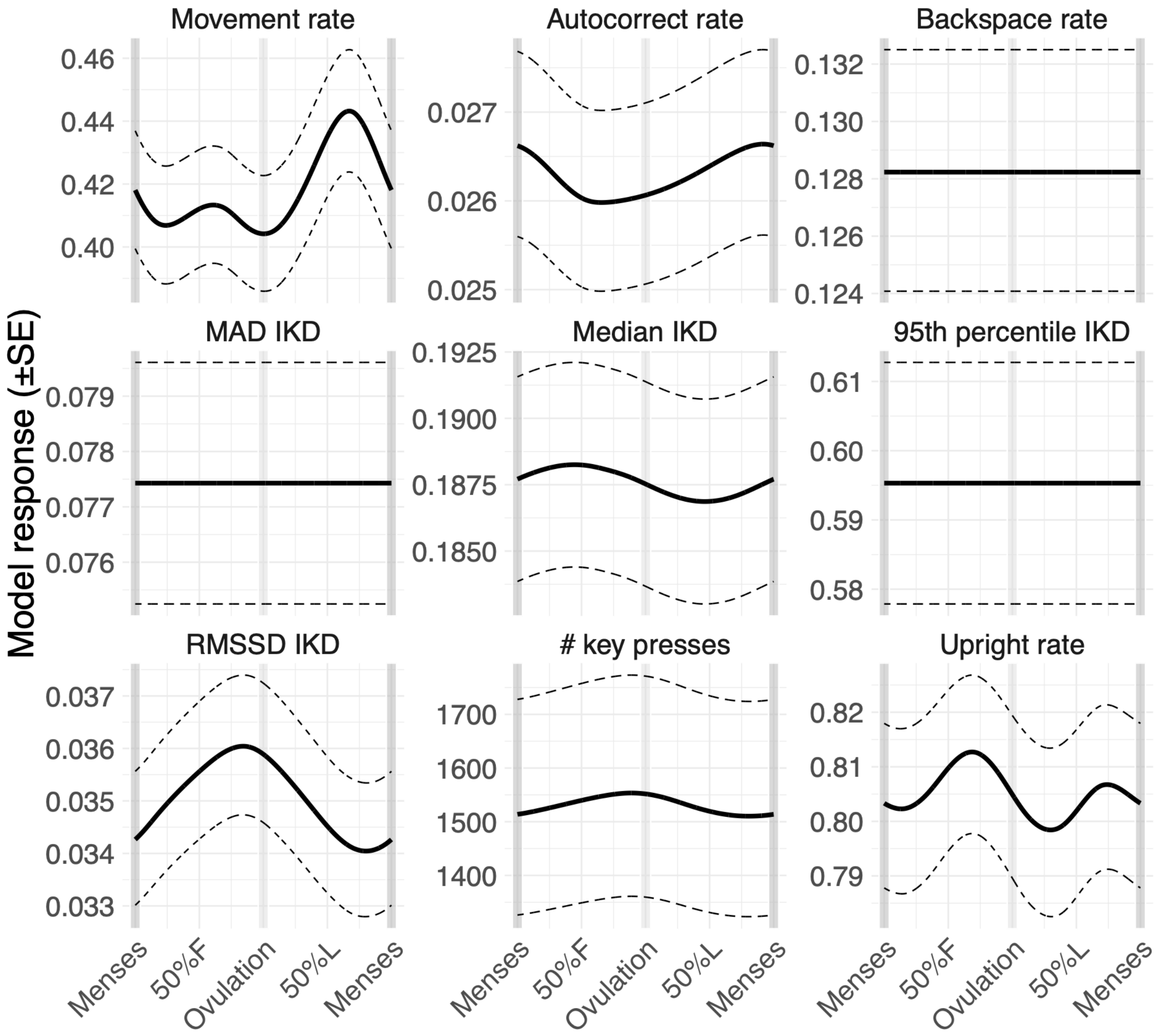

***Supplementary Figure 4: Baseline models centred on ovulation rather than menses onset.*** *IKD = Inter-key delay; MAD = Median absolute deviation; RMSSD = Root-mean-square of successive differences.*

As suggested by Nagpal et al.[1], we reran our baseline analysis on ovulation-centred cycle time to gain a comprehensive picture of typing dynamics across the menstrual cycle. Model structures (response distributions, spline configuration, and random effects) are the same as in the main text.

Overall, results are comparable to the menses-centred baseline models. Movement rate (edf = 3.87, F = 9.29, p = 0.0016), median IKD (edf = 1.57, F = 17.47, p = 0.017), and log RMSSD IKD (edf = 1.68, F = 5.33, p = 0.037) models show significant cycle effects. The cycle effect on autocorrect rate, however, is only marginally significant (edf = 1.9, F = 2.48, p = 0.070). Model response curve shapes are consistent with those of the menses-centred models, except for upright rate. Despite its high wiggliness, the cycle effect was not significant for upright rate (edf = 2.82, F = 1.35, p = 0.22). Estimates and statistics for all other effects are given in Supplementary Table 4.

***Supplementary Table 4***

*Model estimates and associated statistics of ovulation-centred baseline GAMMs. F is a weighted sum of chi-squared statistics. edf = Effective degrees of freedom.*

| Response | Cycle effect | Random intercept | Random cycle slope |
|---|---|---|---|
| Movement rate | edf = 3.87, F = 9.29, p = 0.0016 | edf = 62.56, F = 49.06, p < 0.0001 | edf = 26.86, F = 2.16, p = 0.0015 |
| Autocorrect rate | edf = 1.9, F = 2.48, p = 0.07 | edf = 61.25, F = 72.24, p < 0.0001 | edf = 25.7, F = 4.61, p = 0.00028 |
| Backspace rate | edf = 0.0011, F = 5.2e-05, p = 0.71 | edf = 61.37, F = 79.08, p < 0.0001 | edf = 26.19, F = 4.11, p = 0.01 |
| MAD IKD | edf = 0.00084, F = 0.00013, p = 0.91 | edf = 62.39, F = 7860.22, p < 0.0001 | edf = 0.0037, F = 0.0031, p = 0.82 |
| Median IKD | edf = 1.57, F = 17.47, p = 0.017 | edf = 63.5, F = 10973.02, p < 0.0001 | edf = 14.02, F = 40.98, p = 0.24 |
| 95th percentile IKD | edf = 0.00012, F = 4.7e-05, p = 0.71 | edf = 61.82, F = 2484.22, p < 0.0001 | edf = 13.49, F = 23.85, p = 0.063 |
| Log RMSSD IKD | edf = 1.68, F = 5.33, p = 0.037 | edf = 53.24, F = 502.44, p < 0.0001 | edf = 0.79, F = 0.82, p = 0.41 |
| # key presses | edf = 0.87, F = 0.32, p = 0.23 | edf = 69.93, F = 68.23, p < 0.0001 | edf = 45.23, F = 5.56, p < 0.0001 |
| Upright rate | edf = 2.82, F = 1.35, p = 0.22 | edf = 61.35, F = 41.28, p < 0.0001 | edf = 19.18, F = 1.41, p = 0.0013 |

### Ovulation-centred mixture model

To further explore the effect of cycle centring, we created ovulation-centred mixture models of BiAffect movement rate. Beyond the alternative centring, these were configured as in the main text.

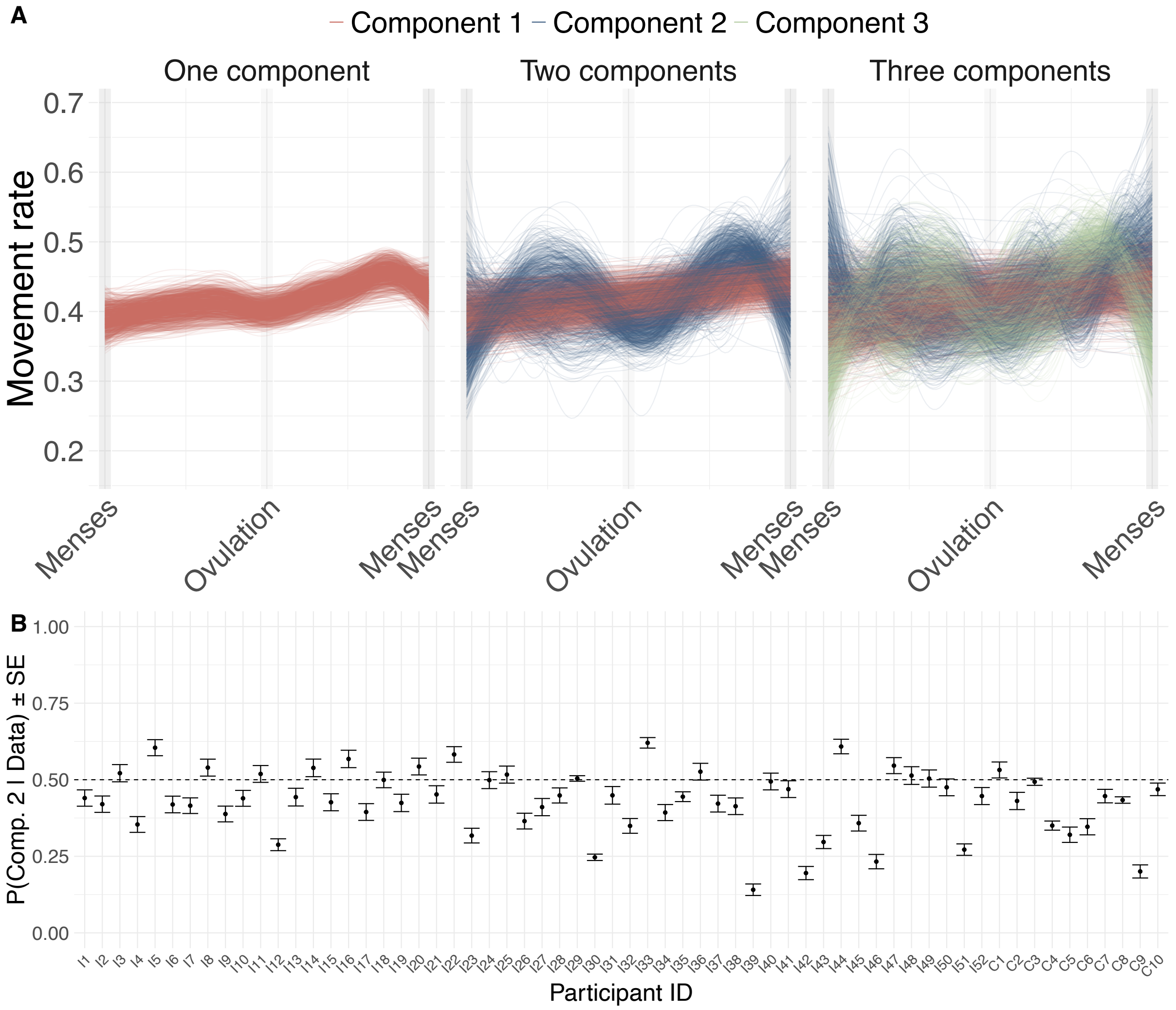

***Supplementary Figure 5: Ovulation-centred mixture models of movement rate and the mean responsibilities of the two-component mixture model. A*** *GAMM curves of component models across all cross-validation iterations.* ***B*** *Mean responsibilities for component 2 of the two-component mixture model (the blue curves in the middle panel of A), calculated from the test sets of every cross-validation iteration.*

Supplementary Figure 5A shows the results of the stability analysis of the ovulation-centred mixture models. The one-component model is largely as expected: the movement rate increases late-luteally, albeit not as much as in the menses-centred mixture models. The two-component mixture model shows larger discrepancies, however. The shape of the second component seems to not only be a shifted but also a smoother, less articulated version of the menses-centred second component, and the secondary increase in the follicular phase seems almost as large as the late-luteal increase. The stability results of the three-component mixture model are as challenging to interpret as in the menses-centred case, but overall it seems that the late-luteal increase shown by component 3 is not very pronounced.

This lack of well-articulated effect might be what drives the pattern we see in Supplementary Figure 5B. The mean component 2 responsibilities of the two-component mixture model are distributed much closer to 0.5 than in the menses-centred case, indicating that the data provide little guidance on which component every individual should be assigned to. All in all, it seems that the ovulation-centred mixture model fails to recover the effects of the cycle on movement rate that we saw in menses-centred mixture models in the main text.

A potential reason for this behaviour is that the effect of interest, namely the late-luteal movement rate increase, occurs at the edge of the domain when we centre on ovulation, and it is not atypical for splines to show suboptimal behaviour at domain edges. This is less of a problem for the ovulation-centred baseline models because they use cyclic cubic splines, which we omitted for the mixture models since we found the cyclic splines could induce extra wiggliness in the flat components despite the high smoothing parameter. To conclude, the late-luteal movement rate effect is best investigated using menses-centred models, which conforms to the recommendations made in Nagpal et al.[1].

#### Lifting the flat component restriction

The two- and three-component mixture models we presented above and in the main text all have the restriction that one of their components is (nearly) flat by forcing their GAMM smoothing parameter to a high value. While this aligns with our a priori hypothesis that some individuals do not show meaningful variation in typing dynamics across the menstrual cycle, it can be insightful to observe the models' behaviour when this restriction is lifted. Therefore, we reran our stability analysis for smartphone movement rate while typing without fixing the component smoothing parameters.

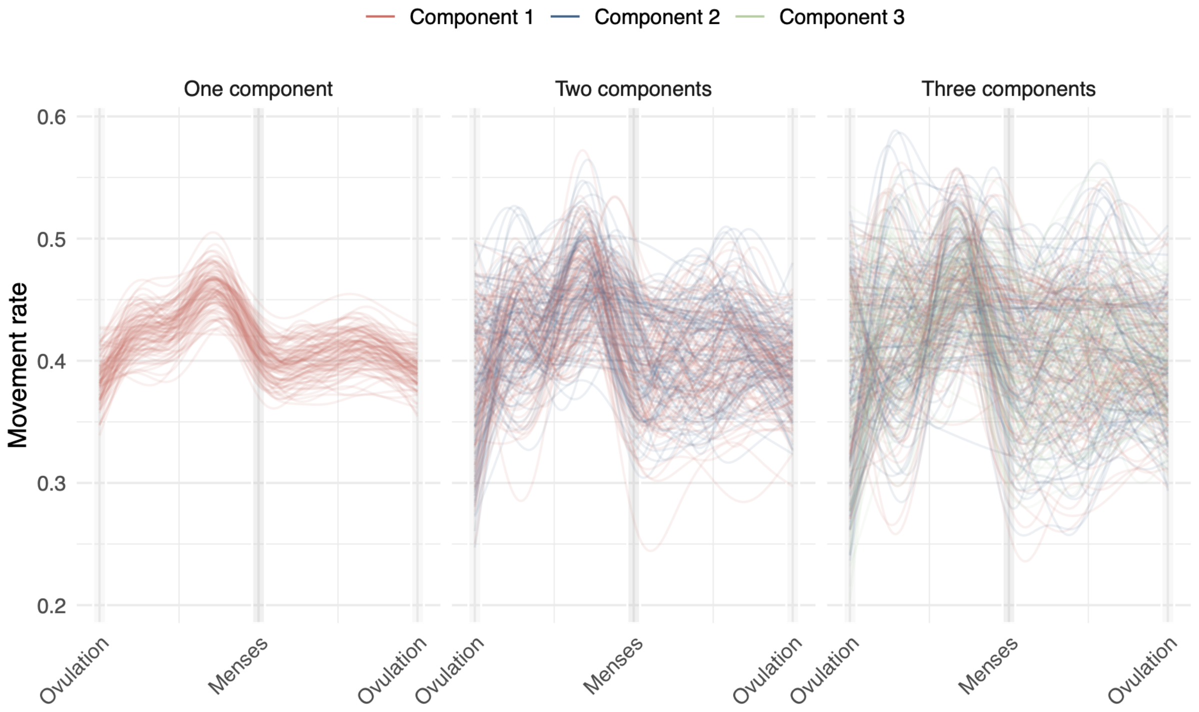

***Supplementary Figure 6: Stability analysis results for unrestricted mixture models of movement rate.*** *Note that component ordering was kept as-is, i.e., we did not use the principal component of the spline coefficients to recolor the component curves.*

To keep the computational cost manageable, we ran the stability analysis with 100 iterations. We used 5-fold cross-validation to find the right set of smoothing parameters in the two- and three-component mixture models. All other algorithm settings were kept the same as in the main text.

Supplementary Figure 6 shows the fitted curves of the unrestricted mixture models. Note that here, we did not use the visualisation trick from the main text (colouring the curves based on the first principal component of the spline parameters), as it is unclear how to generalise that trick to three types of curves. Focusing on the two-component mixture models, however, a visual examination of the individual curves (not shown here) suggests that there is rather high variability in the types of curves that are being fitted. In some cases, components are fitted as a flat line despite being unrestricted, but on many other iterations both components of the two-component mixture exhibit high wiggliness.

Whilst these results could be interpreted as an indication that the data do not always provide sufficient evidence for a flat component, they also demonstrate how restricting the solution space can lead to considerably more stable results. Finding reliable subtrajectories in a medium-sized data set with highly flexible models (of which GAMMs are an example) is a difficult problem that might lead to spurious results if not using proper regularisation. This is most likely why the second component of the restricted two-component mixture model does show very similar patterns across iterations (Figure 2D in the main text), greatly facilitating interpretation and downstream modelling.

### Correcting for physical pain

Many of the symptoms recorded through the daily ratings could be caused, in part, by physical pain experienced during menstruation. To test whether the effects that we find in the daily rating models remain when controlling for pain, we included ratings of physical pain as a parametric (linear) effect in the GAMMs. The pain ratings were centred on the sample mean and scaled to a standard deviation of 1 to aid model fitting. The model equation was updated as follows:

$$g(\mu_{ij}) = \gamma_0 + \gamma_1 m_i + \gamma_2 p_{ij} + b_{i0} + C_{ij} b_{i1} + f_1(C_{ij}) + f_2(C_{ij}) m_i,$$

where the variables are as described as in the Methods but with the addition of the sample-centred pain rating for participant $i$, observation $j$.

In brief, while pain was a significant predictor of all symptoms, including it did not meaningfully alter the cycle and typing subtrajectory membership effects found in the main text (Supplementary Table 5).

***Supplementary Table 5***

*Model estimates and statistics of daily rating GAMMs which include pain as a covariate. Note that all models except Anxiety use inverse link functions, which means that a negative β parameter corresponds to a positive effect. F is a weighted sum of chi-squared statistics. edf = Effective degrees of freedom.*

| | Depression | Anxiety | Anhedonia | Suicidal ideation | Irritability |
|---|---|---|---|---|---|
| $\gamma_1$ (wiggly intercept) | β = 0.099, t(1) = 1.6, p = 0.11 | β = -0.1, t(1) = -0.9, p = 0.37 | β = 0.08, t(1) = 1.22, p = 0.22 | β = 0.055, t(1) = 1.12, p = 0.26 | β = 0.043, t(1) = 0.85, p = 0.4 |
| $\gamma_2$ (pain) | β = -0.01, t(1) = -5.44, p < 0.0001 | β = 0.023, t(1) = 5.34, p < 0.0001 | β = -0.018, t(1) = -7.23, p < 0.0001 | β = -0.016, t(1) = -7.27, p < 0.0001 | β = -0.019, t(1) = -6.88, p < 0.0001 |
| $f_1(C_{ij})$ (cycle) | edf = 1.05, F = 0.46, p = 0.22 | edf = 3.09, F = 9.43, p = 0.00051 | edf = 0.0032, F = 5.9e-05, p = 0.92 | edf = 3.76, F = 6.93, p = 0.0015 | edf = 3.16, F = 4.03, p = 0.00011 |
| $f_2(C_{ij})$ (wiggly cycle difference) | edf = 3.17, F = 8.9, p < 0.0001 | edf = 0.0062, F = 1e-04, p = 0.94 | edf = 3.89, F = 6.64, p = 2e-04 | edf = 0.16, F = 0.02, p = 0.38 | edf = 0.73, F = 0.12, p = 0.31 |
| $b_{i0}$ (random intercepts) | edf = 55.84, F = 54.62, p < 0.0001 | edf = 55.14, F = 37.6, p < 0.0001 | edf = 55.03, F = 46.9, p < 0.0001 | edf = 55.48, F = 59.79, p < 0.0001 | edf = 53.08, F = 19.09, p < 0.0001 |
| $b_{i1}$ (random slopes) | edf = 28.95, F = 5.6, p = 0.0074 | edf = 28.1, F = 7.12, p < 0.0001 | edf = 21.39, F = 1.79, p = 0.15 | edf = 32.55, F = 3.64, p = 0.00015 | edf = 18.95, F = 0.85, p = 0.22 |

### Daily rating model diagnostics

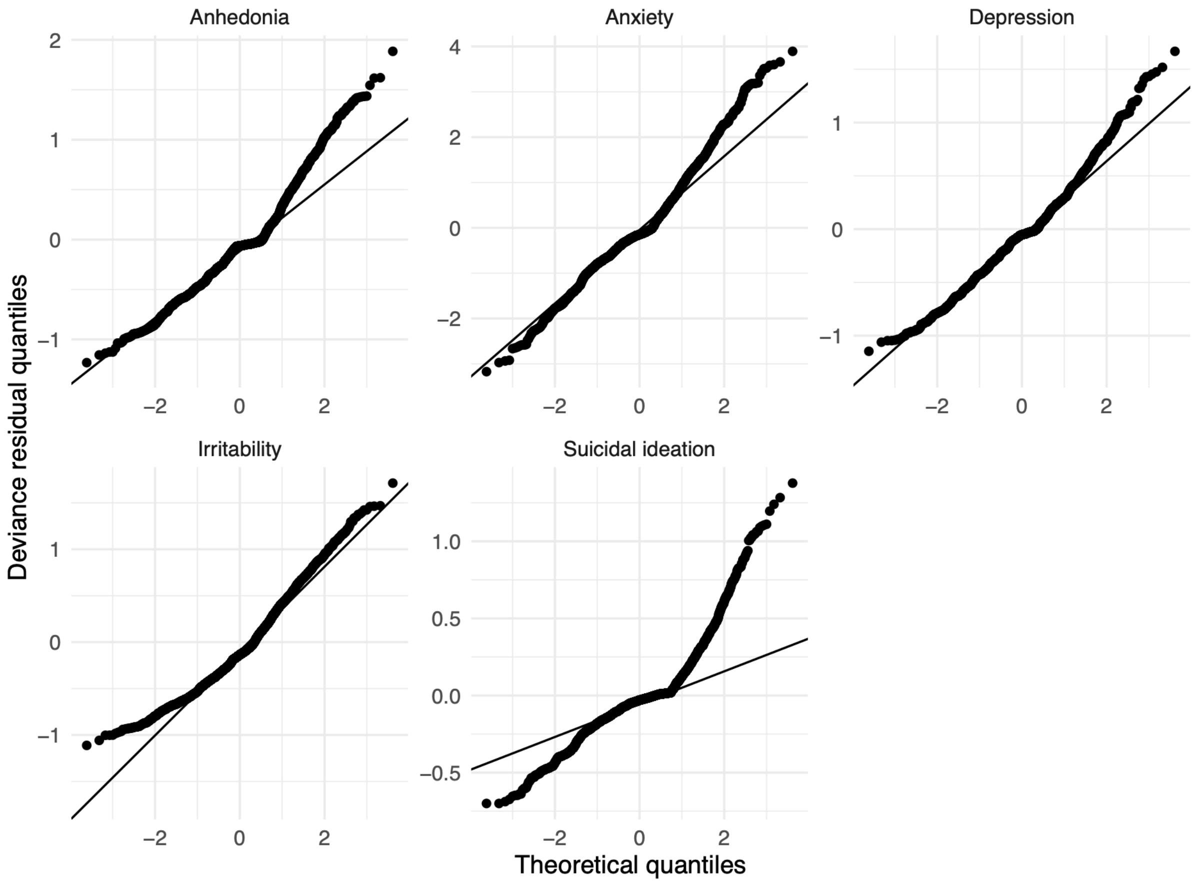

***Supplementary Figure 7: Quantile-quantile plots of the deviance residuals of the daily rating GAMMs.***

### Daily rating items

The depression ratings consisted of 6-point Likert scale responses to the statement "Felt depressed, down, or blue".

The anhedonia ratings consisted of 6-point Likert scale responses to the statement "Did not enjoy my usual activities (e.g., work, school, friends, hobbies)".

The irritability ratings consisted of 6-point Likert scale responses to the statement "Felt irritable".

The anxiety ratings consisted of 6-point Likert scale responses to the statement "Felt anxious, keyed up, or on edge".

The suicidal ideation score was a composite consisting of the mean of several ratings of items from the Adult Suicidal Ideation Questionnaire (ASIQ) and some miscellaneous items. More specifically, the items asked participants to rate the following statements on a 5-point Likert scale: 1) "I wished I were dead"; 2) "I thought that life was not worth living"; 3) "I wished I could go to sleep and not wake up"; 4) "I thought it would be better if I was not alive"; 5) "I thought about killing myself"; 6) "I thought about killing myself, but would not do it"; 7) "I thought that if things would not go better I would kill myself"; 8) "I wanted to kill myself".

### Continuous responsibilities

Before the movement rate mixture model responsibilities (i.e., the component 2 membership) as moderators in the daily rating models, we binarised them. One could also keep them as continuous values between 0 and 1 and fit an interaction surface between them and the PACTS values. The model equation then becomes:

$$g(\mu_{ij}) = \gamma_0 + b_{i0} + C_{ij}b_{i1} + f_1(C_{ij}) + f_2(r_i) + f_3(C_{ij}, r_i),$$

where the parametric and cycle difference effects of the binarised subtrajectory membership, $m_i$, have been replaced with a main smooth effect and a two-way smooth interaction effect of the continuous responsibility $r_i$ for the second component of the two-component mixture model.

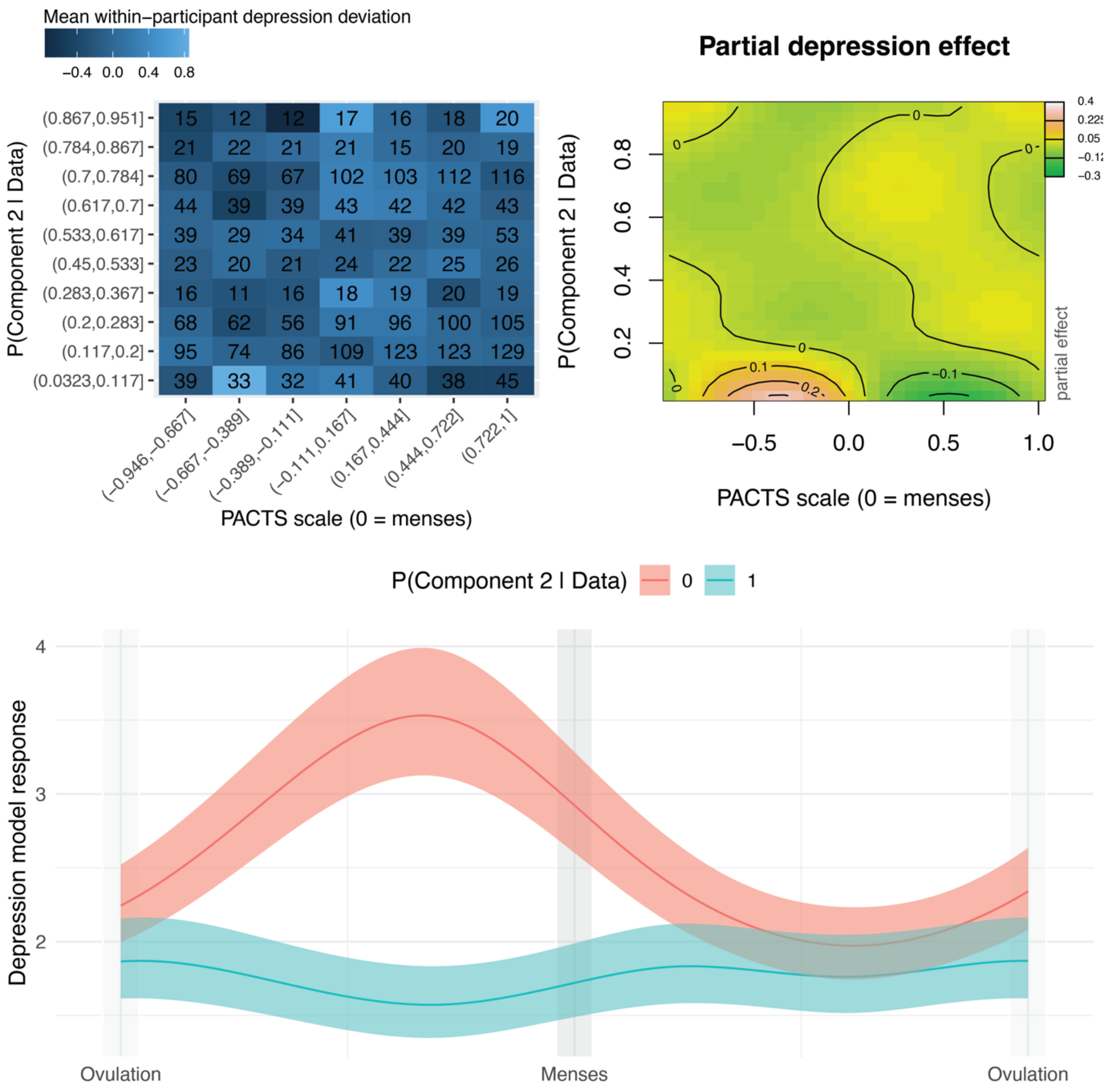

***Supplementary Figure 8: Partial and summed effects of an interaction between cycle time and the continuous component 2 responsibilities.*** *Top-left: participant-centred depression rating averages (colour), calculated for binned values of PACTS values (x-axis) and responsibilities (y-axis). Numbers within the cells indicate the number of observations. Top-right: partial effect of the two-way interaction between responsibilities and PACTS values. Lighter colours denote higher values. Bottom: summed effect for a responsibility of 0 and of 1.*

We visualised the effects of such a model for depression ratings in Supplementary Figure 8. The bottom panel shows patterns that do not match the daily rating results reported in the main text, in that depression fluctuates more for low responsibilities (indicating a higher probability of belonging to the flat movement rate group) than for high responsibilities (higher probability of beloning to the wiggly movement rate group). The partial interaction effect (top-right panel) shows that these fluctuations are confined to low responsibility values, with a large late-luteal peak. Responsibility values above 0.2, on the other hand, are much more stable.

The origin of this effect can be understood by considering the top-left panel of Supplementary Figure 8, which shows a matrix of participant-centred depression averages calculated per cycle and responsibility bin. One cell in the bottom-left of the matrix shows a particularly high depression rating whilst only consisting of 33 observations, and this seems to be the primary source of the interaction effects. Although this does not necessarily mean that the effect is “fake”, we should not overinterpret it based on such a limited number of observations.
